# Decomposing genetic effects of social connectedness and depression on youth behaviors and long-term clinical outcomes

**DOI:** 10.1101/2025.11.21.25340781

**Authors:** Lucy Shao, Shreya Pakala, Bohan Xu, Robert Loughnan, Johnathan Ahern, Haixia Zheng, Wesley K. Thompson, Martin Paulus, Chun Chieh Fan

**Author notes:** Corresponding author: Chun Chieh Fan.

## Abstract

**Importance:** Lacking social connectedness is associated with broad morbidity. Yet the underlying mechanisms remain obscures, especially given its entanglement with depressive symptoms. Clarifying whether social connectedness confers unique risk beyond depression can guide clinical care and social infrastructure interventions.

**Objective:** To decompose shared genetic architecture between social connectedness and depressive affect, derive independent polygenic liabilities, and test their differential associations with youth behaviors, digital engagement, and adult clinical outcomes.

**Design, Setting, and Participants:** Using summary statisitcs from eight genome-wide association studies (GWAS) on social connectedness related traits, we extracted two orthogonal latent genetic factors: a depression related factor (DEP) and a social disconnection factor (SOC). We performed GWAS-by-subtraction to obtain single nucleotide polymorphisms (SNP) level effect sizes and then constructed factor specific polygenic scores (PGS) in (i) a multi-site longitudinal youth cohort (Adolescent Brain Cognitive Development Study, ABCD, questionnaire set n=11,378; EARS smartphone subsample n=914) and (ii) a large scale electronic health record adult cohort (All of Us Research Program, AoU, n=402,944). We evaluated their relative contributions to youth’s psychopathology, prosocial behaviors, screen time use, and adult’s clinical outcomes.

**Results:** In GWAS-by-subtraction, SOC has a GWAS significant loci on *TCF4*; DEP has two loci on *TMEM161B* and *OLFM4*. In youth, a 1-SD increase in SOC PGS was associated with lower prosocial behavior (“being nice to others”; parent-report β=–0.11, SE=0.02, P=7×10 ; youth self-report β=–0.07, SE=0.01, P=4×10) and higher externalizing problems (“argues a lot”; β=0.12, SE=0.02, P=8×10 ¹). In contrast, a 1-SD increase in DEP PGS was more strongly associated with internalizing psychopathology (“too fearful or anxious”; β=0.15, SE=0.02, P=2×10 ¹ ; “unhappy, sad, or depressed”; β=0.14, SE=0.02, P=1×10 ¹) and with fewer high-functioning peers (“friends are athletes”; β=–0.12, SE=0.02, P=3×10 ¹¹; “friends are excellent students”; β=–0.08, SE=0.01, P=4×10 ¹¹); effects replicated directionally in non-European youth. In the EARS subsample, higher SOC and DEP PGS were each associated with greater smartphone screen time. In AoU, SOC PGS was significantly associated with 185 of 712 phecodes (26.0%) and DEP PGS with 167 of 712 (23.5%).

**Conclusions and Relevance:** Social connectedness and depressive affect are highly correlated yet genetically separable with distinct behavioral and clinical profiles. Addressing both may be necessary to reduce the health burden associated with the rising reports on loneliness.

## Introduction

Social connectedness is an individual state characterized by subjective perception on being included, actively prosocial engagement, and synchronized interpersonal alignment ^1,2^. Despite its critical function in human society, the perceived lack of social connectedness is prevalent and on the rise. In 2022, one in three adults in the United States reported to be lonely and one in four reported lacks social support ^3,4^. The “epidemic of loneliness” ^5^ were cooccurring with cardiometabolic disease, psychiatric disorders and premature mortality ^4^. Those observational findings were consistent with results from genetic studies, where genetic tendencies toward loneliness were found to be associated with increasing risks in broad spectrum of physical disorders ^6–8^. Since social infrastructure can be modified to enhance social support and social engagement ^9–11^, improving the social connectedness might be a readily deployable strategy to improve the long-term well-being and physical health in scale.

However, the etiological link between social connectedness and health outcomes is far from clear. Individuals with depression perceive loneliness more often and would have evident reduced social activities when symptoms exacerbated ^12–14^. Given the detrimental effects of depression on broad range of health outcomes, it is unclear if the associations between self-report loneliness and clinical outcomes were good indicators for the clinical relevance of social connectedness . Indeed, a study using data from UKBiobank showed that majority of the associations between social disconnection, measured by self-report loneliness, and clinical outcomes can be explained away by the existence of depressive symptoms ^15^. With the surging prevalence of depressive disorders in recent years ^16,17^, one might wonder if focusing on social connectedness might be diverting the resource away from treating depressive symptoms.

This dilemma can be further exemplified by the social media use among youth. Increasing screen time is associated with perceived loneliness and risks of depression among youth ^18–20^. However, it can be that depressive individuals have increased screen time on the phone due to social withdrawal. Increased screen time can also be driven by prosocial tendencies among youth to seek out alternative ways of social connections. Yet, prior studies did not have instrumental variables that separate the sensitivity to perceived loneliness from the prosocial tendencies. Both large scale observational and genetic studies on social connectedness rely on self-report subject feelings, such as feeling lonely, that is closely related to depressive tendencies ^7,21,22^. The active component of the prosocial engagement, such as tendencies to seek out social connections or participate social activities, were less explored ^6^.

To clarify the relationships between prosocial engagement and depressive affect, here we explicitly decomposing the genetic effects of social connectedness into two distinct and independent latent factors, using eight summary statistics from genome wide association studies (GWAS). One for the subjective feelings on loneliness that is closely related to depression (DEP) and another for the tendencies in engaging social activities that represents the salient risks in social disconnection (SOC). After examining their genetic architecture, we applied the derived GWAS weights to calculate their corresponding polygenic scores (PGS) in two large scale genetic studies. In Adolescent Brain Cognitive Development Study (ABCD, n = 11,378), we investigate the relative contributions of SOC PGS and DEP PGS on youth’s psychopathologies and screen time use. In All of Us Research Program (AoU, n = 402,944) ^23^, we examined the associations of SOC PGS and DEP PGS with broad spectrum of clinical outcomes. By comprehensively studying the non-overlapping genetic signals, we aim to re-evaluate the relative contributions of social connectedness to health, and to clarify whether the risk of social disconnection, after partializing out the depressive affect component, still represents a distinct, genetically grounded pathway through which strengthening social infrastructure might improve population health.

## Material and Methods

### Estimating genetic effects on two independent latent factors

We selected eight different traits that have been used to examine the social connectedness related construct in the context of GWAS. Details of those eight GWAS summary statistics can be found in **Table S1**, including six from 23andMe ^24–27^ (Recharge by socializing, feel left out, physical activity, life satisfaction, loneliness, and self-report depression), one from multi-variate composite measure of social interactions in UKBiobank ^6^, and one from depression diagnoses by Psychiatric Genetic Consortium ^28^. We adopt the strategy called GWAS-by-subtraction ^29^, applying their model with GenomicSEM ^30^ to establish two independent latent factor from eight GWAS summary statistics (See **Figure 1A to 1B**). We then perform GWAS on those two independent latent factors, using the same procedure described in ^29^. The corresponding GWAS results are summarized with FUMA ^31^, and fed to PRS-CS ^32^ to obtain posterior weights for subsequent PGS calculations.

**Figure 1.**
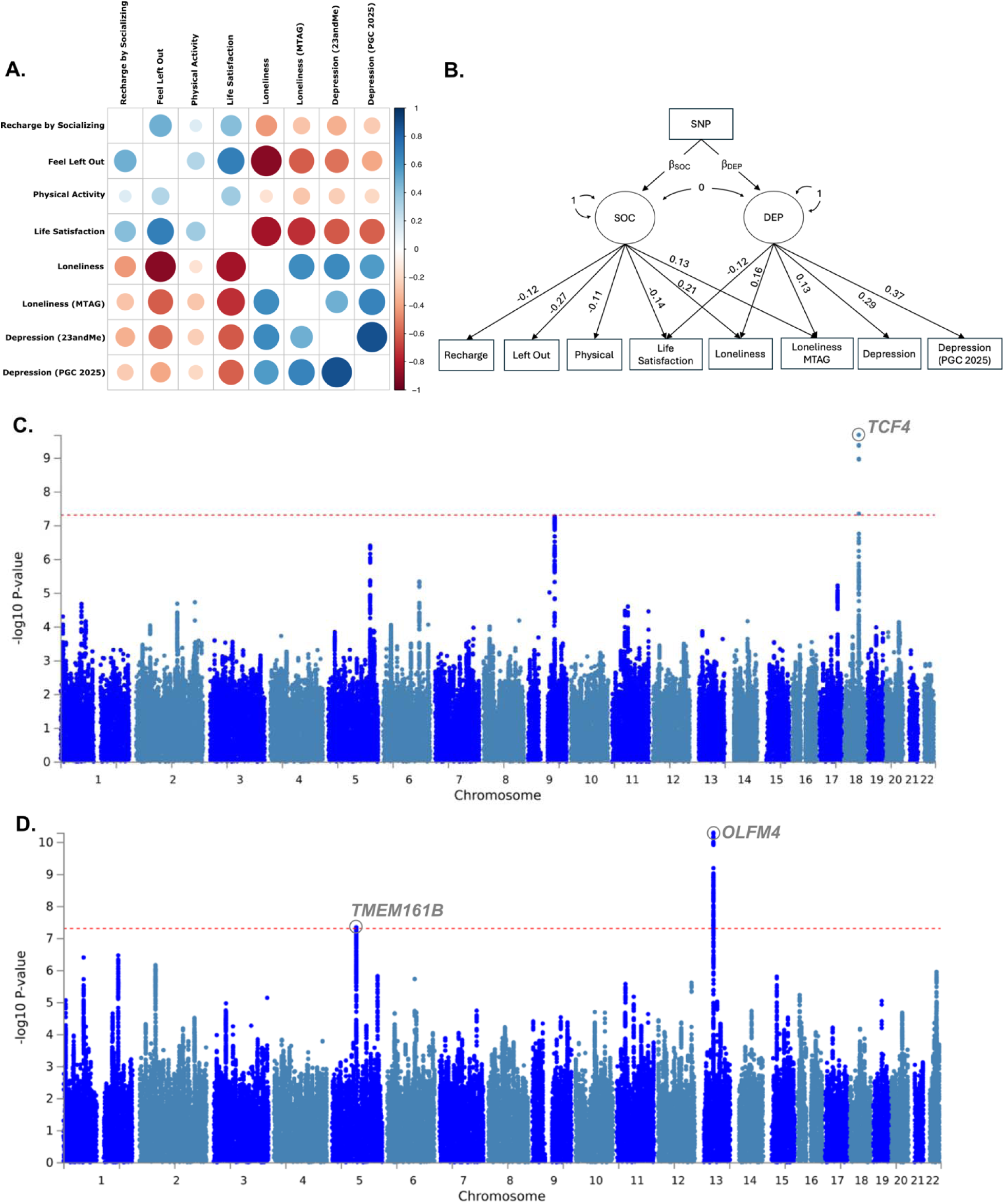
GWAS-by-subtraction for social connectedness. (A) Genetic correlations across eight GWAS, estimated using LDSC. The coloring and size of each cell represent the estimated effect sizes. (B) GWAS-by-subtraction model. Two latent factors were forced to be orthogonal, SOC to represent the lacking of prosocial engagement and DEP to represent the depressive affect. Traits were order listed according to previous panel. The parameter estimated between latent factors and observed GWAS were labeled. (C) Manhattan plot on GWAS of the SOC latent factor. (D) Manhattan plot on GWAS of the DEP latent factor.

### PGS associations in ABCD Study

This study uses ABCD Study data release 6.0. The ABCD Study–the largest in the U.S. assessing longitudinal brain development–was initiated by the United States National Institutes of Health (NIH) in 2015. Starting aged 9-10 years, children are being followed annually with comprehensive measures, including both self- and parent-report questionnaires ^33,34^. More details about the ABCD Study can be found at http://abcdstudy.org.

In the first set of analyses on ABCD Study, we focus on the psychopathologies and social behaviors among youth, based on self-report questionnaires. Detailed questionnaire items can be found in **Table S2**. Of 233 items tested, there are six items from Prosocial Behaviors (PSB) ^35^, 108 items from Child Behavior Checklist (CBCL) ^36^, 101 items from Youth Self-Report (YSR) ^36^, five items from Parent Monitoring Questionnaire (PM) ^35^, six from Peer Behavior Profile (PBP) ^35^, and six items from Family Environment Scale (FES) ^35^. Those questionnaires have been administrated in annual basis for all participants, resulting in a rich dataset with repeated measures. Given that the polygenic score weights are derived from European population, we divided ABCD data into discovery set (n = 6,691) who are genetically similar to continental Europeans (ABCD discovery, EUR set), and replication set (n = 4,687) who are genetically dissimilar to continental Europeans (ABCD replication, non EUR set).

We used imputed genotype data of ABCD Study to calculate PGS. Details of the genetic data and the corresponding quality controls can be found elsewhere ^37^. PGS is based on the weighted sum between posterior weights of GWAS on latent factors and allelic dosages from the quality controlled genetic variants ^38–40^. After deriving social disconnection polygenic score (SOC PGS) and depression polygenic score (DEP PGS) in ABCD Study, we investigate their associations with questionnaire items using cumulative link mixed models (*ordinal* in R ^41^), controlling for repeated measures, family relatedness, first 10 genetic principal components, sex, and age-at-assessment. Both SOC PGS and DEP PGS are in the same model to ensure the estimated effects are independent. Multiple comparisons were corrected with Bonferroni correction.

In the second set of analyses, we use data from those who have participated Effortless Assessment of Risk States (EARS) which recorded patterns of mobile phone usage ^20^. While the study samples reduced to 914 individuals with up to two time-point data, we use this unique dataset to examine the relationships between social disconnection and social media use. Details of each screen time related items from EARS can be found in **Table S3**. Linear mixed effects models are used here, with both SOC PGS and DEP PGS in the same model while controlling for repeated measures, first 10 genetic principal components, sex, and age-at-assessment. Multiple comparisons were corrected with False Discovery Rate.

### PGS associations in AoU

The cohort consisted of participants in the v8 release of the AoU Research Program. The AoU dataset includes electronic health records (EHR), whole genome data, physical measurements, and health questionnaires. The AoU data have been described previously ^42^. Participants were excluded if: (1) they indicated their sex at birth as *other*, (2) they live in a U.S. territory, (3) their state value was missing, (4) they had no EHR data, or (5) they did not have any genomic data. The final sample consisted of 402,944 participants in total.

We then separated the included samples into three independent and mutually exclusive cohorts, (1) participants who have short-read whole genome sequencing (WGS) and are genetically similar to persons of European ancestry (Discovery set, n = 228,064), (2) participants who have WGS and are genetically different than European ancestry (Replication set, n = 174,880). PGS are calculated by applying the posterior effect sizes to the Allele Count/Allele Frequency (ACAF)-thresholded short-read sequencing data provided by AoU. This data was filtered based on a preset ACAF threshold, which required that either the population-specific allele frequency (AF) exceeded 1% or the population-specific allele count (AC) was greater than 100 in any of the ancestry subpopulations. We then excluded sites based on four criteria: (a) excess heterozygosity, (b) overall AF of 0.5% or less, (c) multi-nucleic alleles, or (d) a call rate under 99%.

To comprehensively examine the health outcomes, we scan through clinical diagnoses, based on the grouping of Phecode 2.0 ^43^, limiting to those that have at-least 2,000 cases in the analytic sets. This eventually leads to 712 unique phecodes, spanning across 17 diagnostic categories (detail list in **Table S4**). In each association, we used Firth regression to estimate the effects of SOC PGS and DEP PGS on the binary diagnostic outcomes, controlling for first 20 genetic principal components, age, and sex. Multiple comparisons are corrected through Bonferroni correction, considering 1,424 independent tests.

## Results

### Estimating genetic effects for independent social disconnection factor and depression factor

Using LD score regression ^44^, implemented in genomicSEM ^30^, we estimated genetic correlations of eight included GWAS summary statistics (**Figure 1A**). All included traits had significantly correlated after correction of multiple comparisons (rg range from =0.90 to 0.88, P_Bonferroni_ < 1e-4; **Table S5**), except the genetic correlation between physical activities and loneliness (rg: -0.16, se: 0.09). The self-report loneliness, regardless the source population (23andMe or UKBiobank), was positively correlated with depression, while negatively correlated with other pro-social related measurements. Based on the estimated genetic correlations, we built a GWAS-by-subtract model ^29^ to extract two latent and independent factors. One represented the lack of active prosocial engagement, as the social disconnection latent factor (SOC) to align with the purported risk direction of loneliness construct. The other represented the depressive affects (DEP), to characterize the depression driven component in the perceived loneliness. The model specifications and the estimated model parameters are shown in the **Figure 1B**. The model imposed independency between two latent factors so that our down-stream analyses can focus on the differential effects of SOC and DEP.

The GWAS results on SOC and DEP are shown in the **Figure 1C** and **Figure 1D**, respectively. SOC latent factor has one GWAS significant loci on the intronic region of *TCF4* (18q12.1, rs613872, P = 2e-10). The regional plot is shown in the **Figure S1, panel A**. Insufficient expression of *TCF4* is the cause of Pitt-Hopkins syndrome, a rare genetic disorder with evident developmental delays and deficits in social interactions ^45,46^. Experimental studies have shown that *TCF4* is crucial to the neurodevelopment on social functioning ^47–49^. DEP, on the other hand, has two GWAS significant loci (5q14.3, rs13190179, P = 4.6e-8; 13q14.3, rs2806933, P = 5e-11). Both were reported in GWAS on depressive symptoms and neuroticism ^50^. rs13190179 is on the gene body of *TMEM161B* (**Figure S1, Panel B**), which is related to neocortical development ^51^. rs2806933 can be mapped on *OLMF4* (**Figure S1, Panel C**), which regulates immune responses and inflammations in human intestines ^52^ and has shown to be a potential biomarker for antidepressant responses ^53^. The results of the top associated loci can be found in **Table S6**.

### Independent genetic correlations to psychopathologies and screen times among youth

With the GWAS-by-subtraction results, we applied the posterior weights of the SNPs on the genotype data of ABCD Study to derive SOC PGS and DEP PGS for individual level analyses in ABCD Study. First, we examined the independent contributions of SOC and DEP to the behavioral profiles of the youth (**Figure 2A**). In the discovery set, SOC is significantly associated with 13 items from externalizing domain of CBCL and YSR, 4 items from internalizing domain of CBCL and YSR, 3 items from other problems, and 5 items from social environment and behavior (PSB, PM, PBP, and FES) (All P_Bonferroni_ < 0.05, two tail; details in **Table S7**). For instance, SOC PGS, as its direction tracking the social disconnection, is negatively associated with prosocial behavior (“being nice to others”, parent report: beta = – 0.11, se = 0.02, P = 7E-8; youth self-report: beta = -0.07, se = 0.01, P = 4e-6) while positively increased with externalizing problems (“argues a lot”, beta = 0.12, se = 0.02, P = 8e-10). DEP PGS, on the other hand, is significantly associated with internalizing psychopathologies (“Too fearful of anxious”, beta = 0.15, se = 0.02, P = 2e-14; “Unhappy, sad, or depressed”, beta = 0.14, se = 0.02, P = 1e-14), while reducing with number of friends individuals reported (“Number of friends are athletes”, beta = -0.12, se = 0.02, P = 3e-11; “Number of friends are excellent students”, beta = -0.08, se = 0.01, P = 4e-11). With a formal test on the effect size differences, DEP PGS shows stronger associations than SOC PGS in three internalizing items while SOC PGS shows evidence in stronger associations in one prosocial item (**Figure 2B**). The associations found in discovery set are well replicated in the replication set despite smaller sample size and mismatched population backgrounds that are known to increase statistical uncertainties in PGS analyses ^40^. The effect sizes are consistent between discovery set and replication set, explaining 89% variance in the estimation by mixed effects meta-analysis (**Figure 2C**).

**Figure 2.**
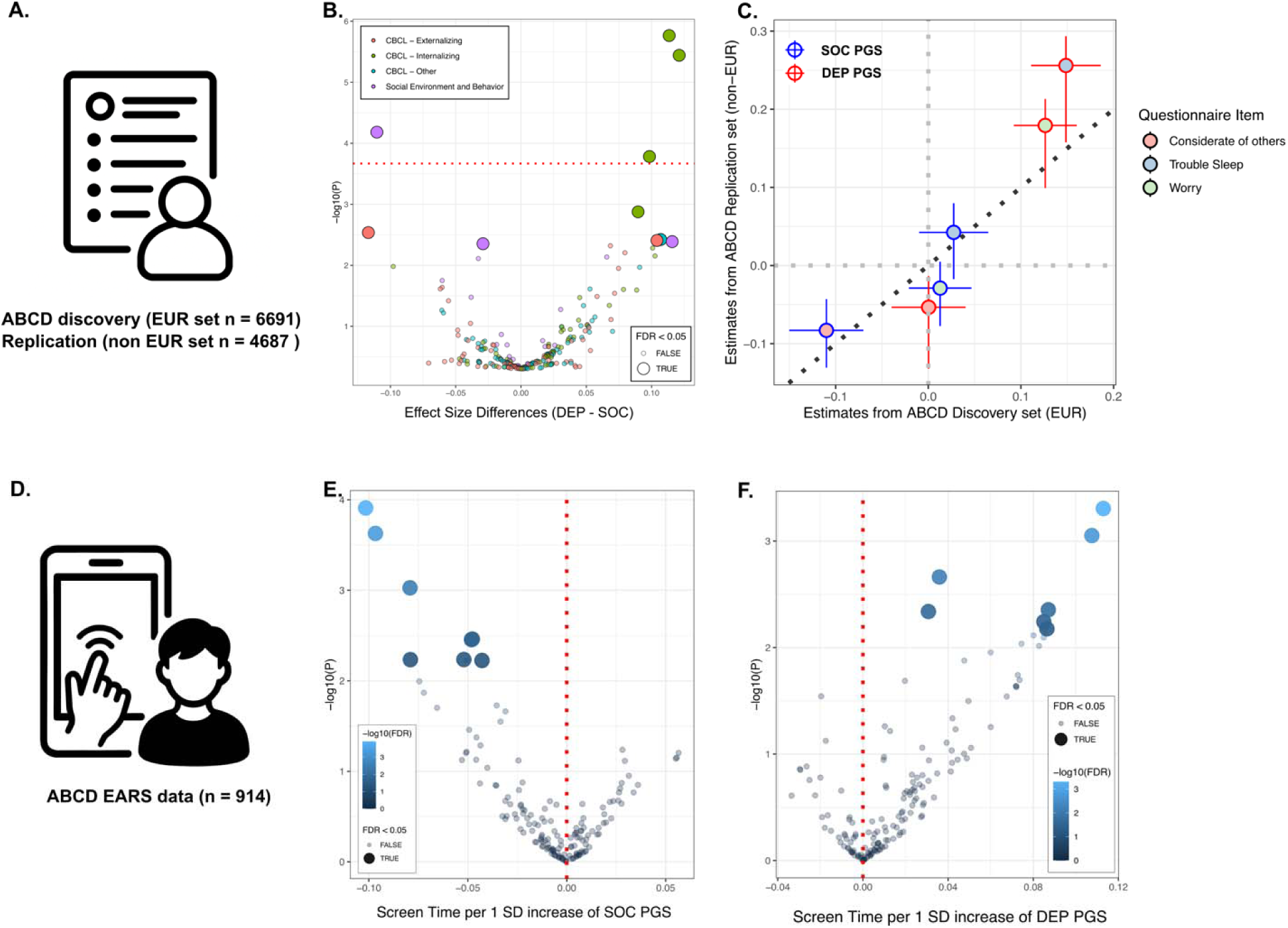
PGS associations with behaviors and screen time use among youth. (A) ABCD samples included in the associations with questionnaire items. (B) Volcano plot on the association differences between SOC and DEP PGS, per 1 SD of the PGS score. (C) Effect size distributions between ABCD discovery and replication. Point fill indicate the questionnaire items while the outline coloring represents which PGS is shown. 95% CI is plot for both discovery and replication. (D) ABCD samples with EARS data. (E) Associations between SOC PGS and screen time measurements. (F) Associations between DEP PGS and screen time measurements.

We then further examine the relationships between mobile phone usage and social disconnection in ABCD Study (**Figure 2D**). SOC PGS is significantly associated with screen time measures from EARS (**Figure 2E**), indicating people with inclinations toward social connections would seek out more interactions on their mobile phones (FDR < 0.05). On the contrary, DEP PGS is positively correlated with screen time among youth (**Figure 2F**, FDR < 0.05), indicating youth with risks of depression also spent more time on their mobile phones. The paradoxical effects from SOC and DEP suggest screen time is a complex end result of social disconnection and depressive symptoms that warrant further in-depth investigations.

### Associations across broad spectrum of clinical outcomes among adults

To investigate the long-term health outcomes given the propensities in social disconnection, we applied the same posterior weights on the WGS data of AoU, deriving SOC PGS and DEP PGS for individual level analysis. We performed Phenome-wide association study (PheWAS), examining the contribution of SOC PGS and DEP PGS to the risks of diseases, defined by Phecode (see **Material and Methods**). Across 712 clinical phecodes tested in the discovery set (n = 228,064), 185 is significantly associated with SOC PGS and 167 is significantly associated with DEP PGS (P_Bonferroni_ < 0.05, two sided; **Figure 3A; Table S8**). Many of the diagnoses in the mental disorders, neurological, and physical symptoms have significant elevated risks by both SOC and DEP, while DEP PGS has several more significant associations with those categories (**Figure 3B**). SOC PGS has more significant associations in digestive, endocrine/metabolic, circulatory, and musculoskeletal categories than DEP PGS (**Figure 3B**). With a formal test for the effect size differences between SOC and DEP, DEP PGS has significantly stronger associations with Bipolar disorder, Major depressive disorder, Generalized anxiety disorder, and Anxiety disorder, than SOC PGS (**Figure 3C**). Meanwhile, SOC PGS has stronger associations in Tobacco use disorder, dental caries, and electrolyte imbalance than DEP PGS (**Figure 3C**). The associations were well replicated in the replication set (n = 174,880). The associations in the replication set are consistent with the effect size estimates from the discovery (mixed effects meta-analytic beta = 0.51, se = 0.05, P = 1e-23), explaining 26 .4% of the variations despite higher uncertainties in the sample compositions (**Figure S2**).

**Figure 3.**
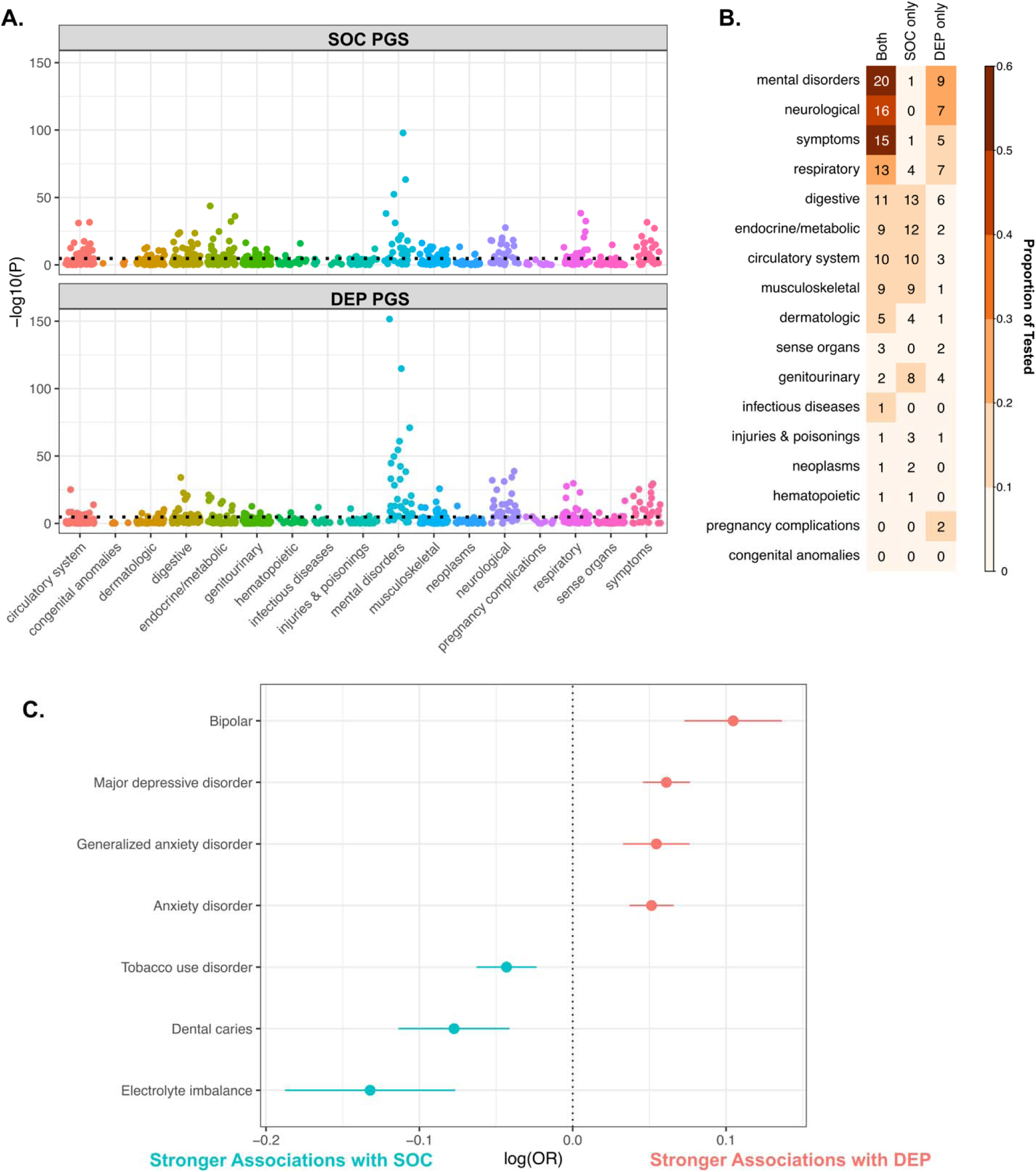
Associations with long-term clinical outcomes. (A) PheWAS on All of Us discovery set (n = 228,064). Each dot represent one phecode while coloring indicate which phecode categories it belongs. Bonferroni corrected significant threshold is plotted as black dotted lines. (B) Counts of significant associations per diagnostic categories. (C) Direct contrasts between SOC PGS and DEP PGS across diagnoses that passed significant threshold. The unit is per 1 SD increase of the differences between to PGS.

## Discussion

We used GWAS by subtraction on eight GWAS summary statistics to extract two independent latent genetic factors to characterize social disconnection (SOC) and subjective loneliness/depressive affect (DEP). Behaviorally, DEP reflects a liability toward internal emotional suffering, e.g. youth high on DEP are described as fearful, anxious, unhappy, and sad, and they tend to have fewer high-functioning peers (fewer friends who are “excellent students” or “athletes”), i.e., they withdraw or struggle socially because they feel bad. In contrast, SOC reflects a liability toward low positive social engagement and conflictual behavior, e.g. youth high on SOC are rated as less prosocial (“less nice/helpful to others”), more argumentative and oppositional (“argues a lot,” “doesn’t follow rules”), and embedded in thinner, more strained social networks, i.e., they are socially disconnected because they do not engage warmly or cooperatively with others, even when overt mood symptoms are less prominent. From genetic loci to polygenic associations, our results showcase the independent and differential signals of SOC and DEP, while presenting a more nuanced picture of their intertwining contributions to youth’s behaviors and adult’s long-term health outcomes. In the loci level, SOC has an unique loci on *TCF4*, a gene implicated in the genesis of autistic behaviors ^46,47,49^, while loci found in DEP are aligning with prior studies on depression and neuroticism ^50^. In the polygenic associations with youth’s behaviors, SOC has evident associations with prosocial behaviors and externalizing problems whereas DEP is more evidently related to internalizing problems. SOC and DEP have divergent effects on the mobile phone usage among youth. Higher disconnection specified by SOC means lower screen time, whereas higher DEP tendency means higher frequencies in using mobile phones. In the level of long-term outcomes among adult, both SOC and DEP independently associated with the risks of broad spectrum of clinical diagnoses. Our results are convergingly indicate that social disconnection and depressive affect are differentiable, contributing to wide range of behavioral and clinical outcomes in a complex and intertwined fashion. Taken together, these converging genetic, behavioral, and clinical findings demonstrate that social disconnection and depressive affect are related but distinguishable liabilities that differentially shape youth social and emotional behavior and confer overlapping yet partly specific risks for adverse health outcomes across the lifespan.

Although the degree of loneliness is the most commonly used measurement for social disconnection, as exemplified in the Morbidity Mortality Weekly Report by Center for Disease Control and Prevention ^4^, it does not capture the complexity of social disconnection and oftentimes conflate with depressive affect. Despite rich text in the epidemiological studies and genetic studies ^3,4,6–8^, there was doubt on how much of the loneliness effects on health can be attributable to social disconnection alone ^15^. Our systematic analyses on the genetic latent factors provide converging evidence to support the unique contribution of social disconnection on behaviors and long-term health outcomes. From the molecular level to population level, SOC and DEP latent factors exhibit differential genetic architecture, both contributing significantly to health outcomes. From the molecular level to the population level, SOC and DEP latent factors exhibit differential genetic architecture, both contributing significantly to health outcomes. Specifically, SOC indexes genetic liability to low prosocial engagement, conflictual and thinned social networks, and reduced participation in mutually supportive relationships that cannot be explained by depressive affect alone, and this liability independently predicts youth externalizing behavior and diverse adult medical morbidities even after accounting for DEP. These findings indicate that social disconnection should be conceptualized, measured, and targeted as a construct distinct from subjective loneliness or depressive mood when designing prevention and intervention strategies. Thus, strengthening social infrastructure and opportunities for positive, stable social engagement—through schools, workplaces, communities, and healthcare systems—may reduce SOC-related risk and yield mental and physical health benefits that cannot be achieved by treating depressive symptoms alone.

Our results further support that role of *TCF4* in social functioning of humans. The loss of function on *TCF4* leads to Pitt-Hopkins syndrome ^45,46^. It has been found to be associated with Autistic behaviors and crucial for neurodevelopment ^47–49^. In our genomicSEM framework, the lead locus at TCF4 loaded specifically on the SOC latent factor, but not on DEP, indicating that TCF4-linked genetic liability is more closely tied to patterns of social engagement, prosocial behavior, and social network quality than to internal emotional suffering or depressive affect.

This pattern is consistent with TCF4’s known role as a transcription factor that regulates neurodevelopmental processes in cortical and hippocampal circuits implicated in social cognition, learning, and flexible behavior, providing a plausible mechanistic bridge between molecular variation and observable social-disconnection phenotypes. Future work should test how TCF4-related liability interacts with environmental exposures—such as opportunities for structured social participation, family and school climate, and community-level social resources—and whether enriched social environments can buffer or redirect the developmental expression of this liability. Taken together, these findings position TCF4 as a key molecular node linking genetic risk for social disconnection to altered social engagement, and they highlight gene–environment interplay in social contexts as a critical target for mechanistic and intervention-focused research.

The differential associations of SOC and DEP among youth provide a more nuanced presentation of behavioral drivers during adolescence. General psychopathologies among youth are determined by multiple factors, intertwining the underlying susceptibility to depression with active social inclinations. It echoes our previous findings that youth’s cognitive functions and general psychopathologies are associated with wide range of polygenic scores, not limiting to specific domains ^38^. This is particularly salient in how youth interacts with the prevalent social outlets, their mobile phones. Socially inclined individuals have more interactions online while people interact online frequently can also have higher genetic tendencies toward depression. Independent and paradoxical contribution of SOC and DEP to the screen time suggests that we cannot rely on simple metrics of screen time to determine the detrimental or beneficial impact of social media use among youth. A more nuanced approach is warranted.

Our study has several limitations. First, discovery GWAS and PGS were derived primarily in European ancestry; although we examined non European replication in ABCD and AoU, cross ancestry attenuation limits generalizability. Second, SOC/DEP depend on the selected input GWAS and GenomicSEM/GWAS by subtraction model specification; alternative input sets could yield slightly different factors. Forced independency between latent factor might not fit the data best in a data-driven point of view, which might even reduce the power for prediction. We focus on the inference for independent effects as that is the key question our study trying to answer. Third, PGS map genetic liability, not causal effects; associations may reflect pleiotropy, gene–environment correlation, or residual confounding.

In light of these caveats, our results reveal that social disconnection (SOC) and depression (DEP) share polygenic liability yet retain separable, clinically meaningful components. SOC is distinguished by a TCF4 signal and a behavioral profile characterized by lower prosocial engagement, higher externalizing and conflictual behaviors, and broader links to cardiometabolic, musculoskeletal, and other physical health outcomes, whereas DEP concentrates genetic risk in internalizing psychopathology and mood/anxiety disorders. These findings indicate that social disconnection is not merely a proxy for depression and that addressing only depressive symptoms will leave an important dimension of risk unmitigated.

Practically, they argue for (i) assessment strategies that capture prosocial behavior, quality and reciprocity of social ties, and social participation in addition to depressive symptoms and loneliness, and (ii) parallel intervention efforts that combine evidence-based treatments for depression with structural and community-level approaches that create stable, rewarding opportunities for social engagement. Ultimately, reducing the health burden associated with today’s era of social disconnection will likely require coordinated clinical and policy initiatives that explicitly target both affective distress and the distinct liability to low, conflictual, or impoverished social connection.

## Data Availability

All data produced in the present study are available upon reasonable request to the authors

## Acknowledgement

This work was partly funded by The William K. Warren Foundation, the National Institute of General Medical Sciences Center (Grant 2 P20GM121312, MPP, CCF), the National Institute on Drug Abuse (U01DA050989, MPP), and the National Institute of Mental Health (R01MH122688, R01MH128959, CCF).

Dr. Paulus advises Spring Care, Inc., receives royalties from an article on methamphetamine in UpToDate, and has a compensated consulting agreement with Boehringer Ingelheim International GmbH.

Data used in the preparation of this article included those obtained from the Adolescent Brain Cognitive Development SM(ABCD) Study (https://abcdstudy.org), held in the NIMH Data Archive (NDA). The ABCD Study® is supported by the National Institutes of Health and additional federal partners under award numbers U01DA041048, U01DA050989, U01DA051016, U01DA041022, U01DA051018, U01DA051037, U01DA050987, U01DA041174, U01DA041106, U01DA041117, U01DA041028, U01DA041134, U01DA050988, U01DA051039, U01DA041156, U01DA041025, U01DA041120, U01DA051038, U01DA041148, U01DA041093, U01DA041089, U24DA041123, U24DA041147. A full list of supporters is available at https://abcdstudy.org/federal-partners.html. A listing of participating sites and a complete listing of the study investigators can be found at https://abcdstudy.org/consortium_members/. ABCD consortium investigators designed and implemented the study and/or provided data but did not necessarily participate in the analysis or writing of this report. This manuscript reflects the views of the authors and may not reflect the opinions or views of the NIH or ABCD consortium investigators. The ABCD data repository grows and changes over time.

We gratefully acknowledge All of Us participants for their contributions, without whom this research would not have been possible. We also thank the National Institutes of Health’s All of Us Research Program for making available the participant data examined in this study.

