## Supplementary Figures for "Decomposing genetic effects of social connectedness and depression on youth behaviors and long-term clinical outcomes"

**Supplementary Materials**

Figure S1. Regional plots for discovered loci from GWAS-by-subtraction

Figure S2. Comparisons on the effect size distributions between All of Us discovery and replication sets.

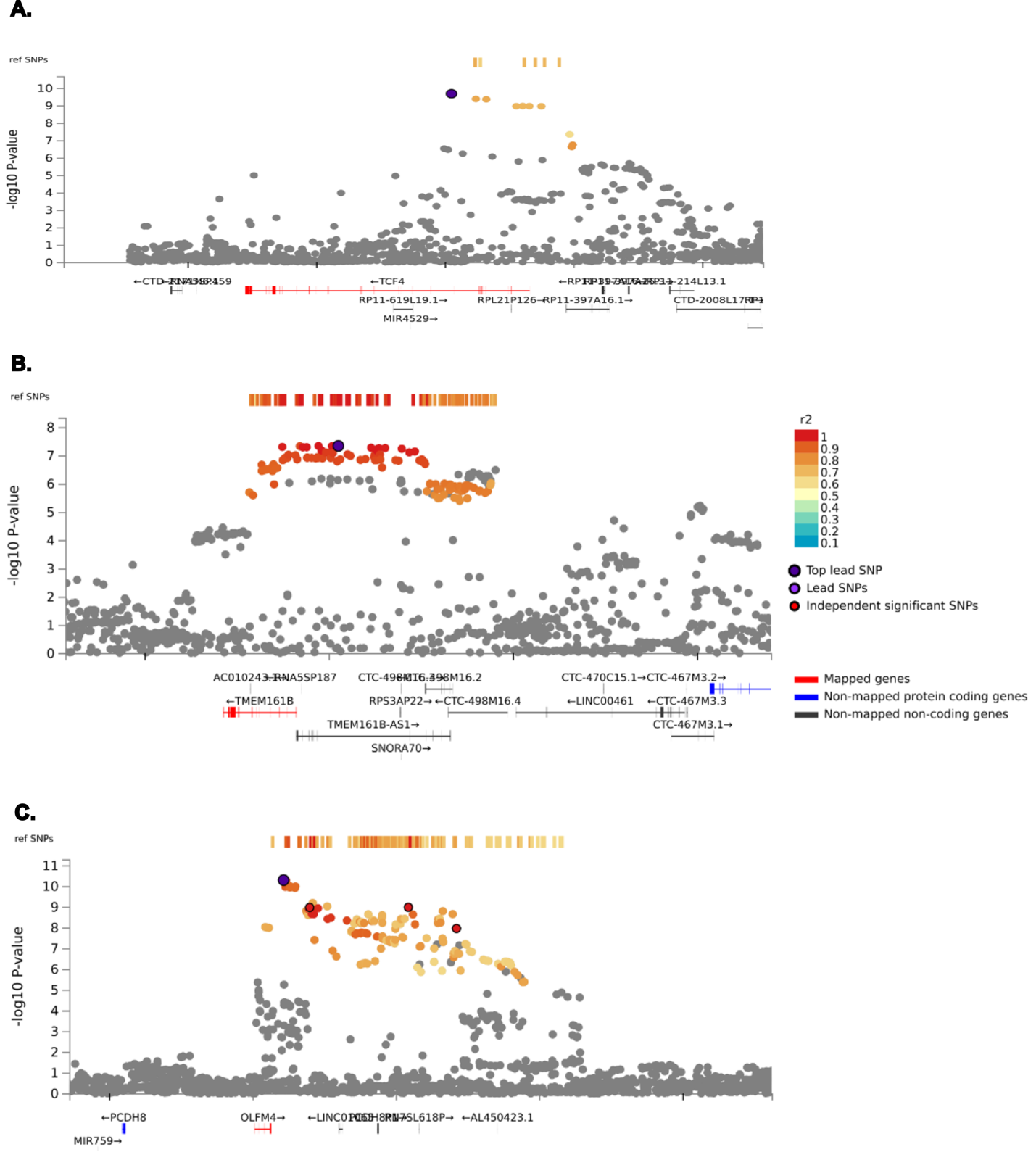

Figure S1.

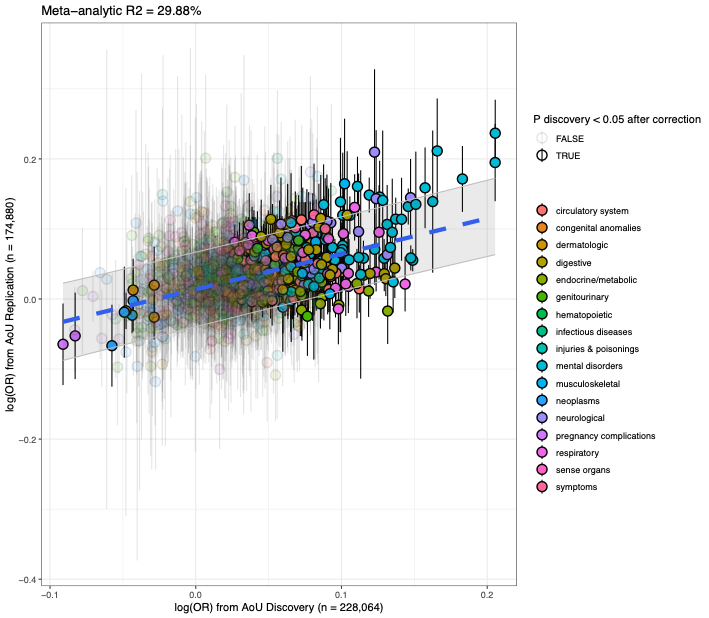

Figure S2
